# Pathogen detection and characterization from throat swabs using unbiased metatranscriptomic analyses

**DOI:** 10.1101/2022.04.08.22273423

**Authors:** Ryan Toma, Nathan Duval, Nan Shen, Pedro J. Torres, Francine R. Camacho, Jiapeng Chen, Oyetunji Ogundijo, Guruduth Banavar, Momchilo Vuyisich

**Author notes:** Corresponding author: Ryan Toma,. 11724 NE 195th St Suite 101, Bothell, WA 98011.

## Abstract

**Objective:** Infectious diseases are common but are not easily or readily diagnosed with current methodologies. This problem is further exacerbated with the constant presence of mutated, emerging, and novel pathogens. One of the most common sites of infection by many pathogens is the human throat. Yet, there is no universal diagnostic test that can distinguish these pathogens. Metatranscriptomic (MT) analysis of the throat represents an important and novel development in infectious disease detection and characterization, as it is able to identify all pathogens in a fully unbiased approach.

**Design:** To test the utility of an MT approach to pathogen detection, throat samples were collected from participants before, during, and after an acute sickness.

**Results:** Clear sickness-associated shifts in pathogenic microorganisms are detected in the participants along with important insights into microbial functions and antimicrobial resistance genes.

**Conclusions:** MT analysis of the throat represents an effective method for the unbiased identification and characterization of pathogens. Since MT data include all microorganisms in the sample, this approach should allow for not only the identification of pathogens, but also an understanding of the effects of the resident throat microbiome in the context of human health and disease.

## Introduction

The human throat contains a large number of both commensal and potentially pathogenic microorganisms performing a wide variety of metabolic functions, with potentially beneficial and harmful effects to the host (Gong et al., 2013). The analysis of throat swabs by metatranscriptomics presents an opportunity to investigate the throat microbiome and its roles in disease pathogenesis.

The throat microbiome has the potential to elucidate several important chronic disease biomarkers. Research has shown that microorganisms in the throat microbiome including the genera *Fusobacterium, Prevotella, Streptococcus, Lactobacilli*, and *Bifidobacterium* are related to specific chronic diseases such as laryngeal carcinoma, Schizophrenia, and cystic fibrosis (Boutin et al., 2015; Castro-Nallar et al., 2015; Gong et al., 2014). In addition to the possible direct effects of the throat microbiome on human health and disease, the throat microbiome can also act as an important proxy to the lung microbiome, which has been shown to have dramatic effects on host physiology (Ibironke et al., 2020). Microorganisms in the lung microbiome (such as *Haemophilus, Neisseria* spp., and *Pseudomonas aeruginosa*) have been associated with numerous diseases, including lung cancer, asthma, and obstructive pulmonary disease (Mao et al., 2018; Moffatt and Cookson, 2017). Since the throat microbiome is easy to sample and can provide informative insights into the lung microbiome, it represents an important development in allowing for large-scale population studies.

In addition to the connections between the throat/lung microbiomes and chronic diseases, the throat microbiome is also an important reservoir for infectious diseases. Throat microbiome sampling can be used to characterize pathogens and can provide insights into infectious disease severity (Lee et al., 2019; Tsang et al., 2019). Current standard clinical practices for the identification of pathogens from throat samples mainly rely either on quantitative real-time PCR (qPCR) or culture assays (Miller et al., 2019). These methods have several drawbacks which have limited the clinical utility of throat swabs and hampered pathogen detection.

Both qPCR and microbial culturing are biased methodologies. These methods rely on the identification of specific target pathogens and are unable to identify novel or unexpected pathogens (Wylezich et al., 2018). Metatranscriptomics represents the ideal method for the detection and characterization of pathogens. Metatranscriptomics can provide strain-level taxonomic resolution for all metabolically active microorganisms and viruses and can provide insights into the biochemical activities of the microbiome by quantifying microbial gene expression levels, allowing for the assessment of biochemical pathway activities (Bashiardes et al., 2016). These analyses are of particular importance to infectious disease diagnostics as they can provide insights into antimicrobial resistance genes to inform treatment options.

To date, metatranscriptomic methods have been limited due to the cost, turnaround time, and complexity of both laboratory and bioinformatic methods (Knight et al., 2012). Effective RNA preservation has been challenging, traditionally requiring a cold chain that is expensive and complicated. Another challenge of working with RNA is that the majority of RNA is uninformative ribosomal RNA (rRNA), with messenger RNA (mRNA), the most informative RNA species, making up only about 1-5% of cellular RNA (He et al., 2010). By removing the non-informative prokaryotic and eukaryotic RNAs, thereby enriching for informative mRNA sequences, sequencing data can be generated at a fraction of the analysis cost (Bashiardes et al., 2016).

Here we present an unbiased method for the quantitative metatranscriptomic analysis of throat samples that can easily be applied to clinical studies, clinical trials, biosurveillance, and clinical testing globally. The method is automated, high-throughput, inexpensive, and includes a fully automated, rapid, and clinically validated bioinformatics suite for strain-level taxonomic classification of all microorganisms, viruses, and their quantitative gene expression. Importantly for infectious disease diagnostics, the current method has a clinically useful turnaround time, and results can be reported within 48 hours of sample collection; the method can be further developed to bring turnaround time to under 24 hours. This level of turnaround time is comparable to traditional methods of infectious disease diagnostics, such as qPCR and cultures (Caliendo et al., 2013; Larremore et al., 2020). An RNA preservation buffer mixed with the throat sample at the point of collection inactivates all pathogens (bacteria, fungi, and viruses), preserves RNA integrity, and enables safe transportation globally at ambient temperatures (Hatch et al., 2019; Toma et al., 2020).

## Methods

### Ethics Statement

All procedures involving human subjects were performed in accordance with the United States ethical standards and approved by a federally-accredited Institutional Review Board (IRB) committee. Informed consent was obtained from the participants, all of whom were over the age of eighteen and residents of the USA at the time.

### Sample Collection

Oropharynx swabs were collected using flocked swabs (Puritan 25-3206-H 20MM). Swabs were collected by sampling across the tonsils and uvula four to five times while avoiding the tongue and cheeks. After collection, swab tips were placed into a bead beating tube containing a proprietary RNA Preservative Buffer (RPB) and broken off. RPB has been previously shown to preserve RNA in clinical samples for up to 28 days at room temperature (Hatch et al., 2019; Toma et al., 2020). Samples were stored at room temperature for up to 2 weeks before being frozen at −80C prior to sample processing.

### Study Design

247 throat swabs were collected from 38 individuals between September 2019 and March 2020 at regular intervals, typically weekly. For participants that naturally developed a sickness, throat swabs were collected at an increased interval, typically daily, until after symptoms subsided, at which point weekly sample collection was resumed.The samples from six participants (male: 4, female: 2) that reported at least one moderate or severe symptom associated with a sickness were analyzed. Throat swabs from prior to the sickness, during the sickness, and after the sickness were analyzed (for a detailed sample collection timeline see supplemental table 1). A symptom survey was administered for each collection to assess if an individual felt sick, and to quantify their symptoms (supplemental table 2).

**Table 1:**
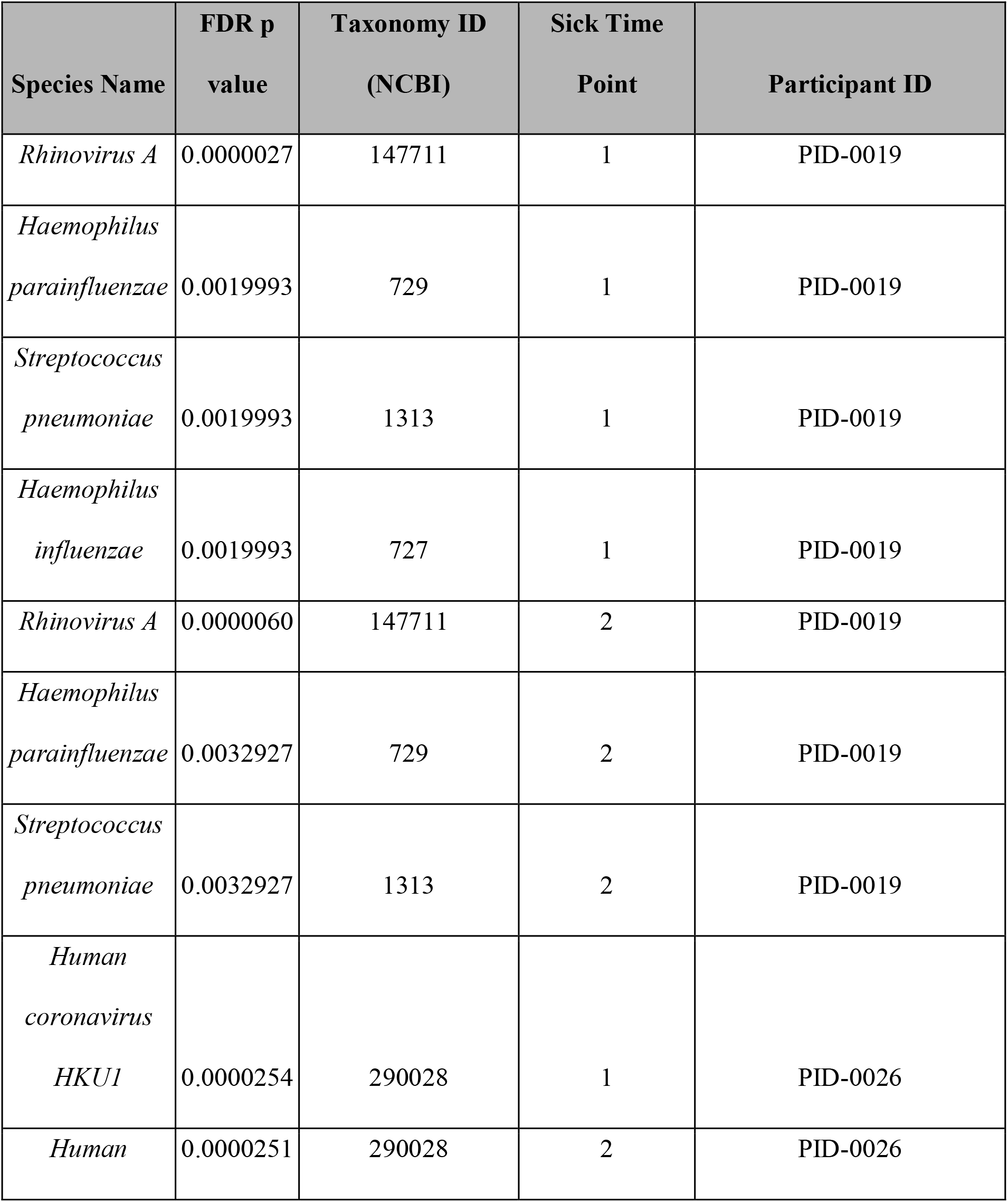
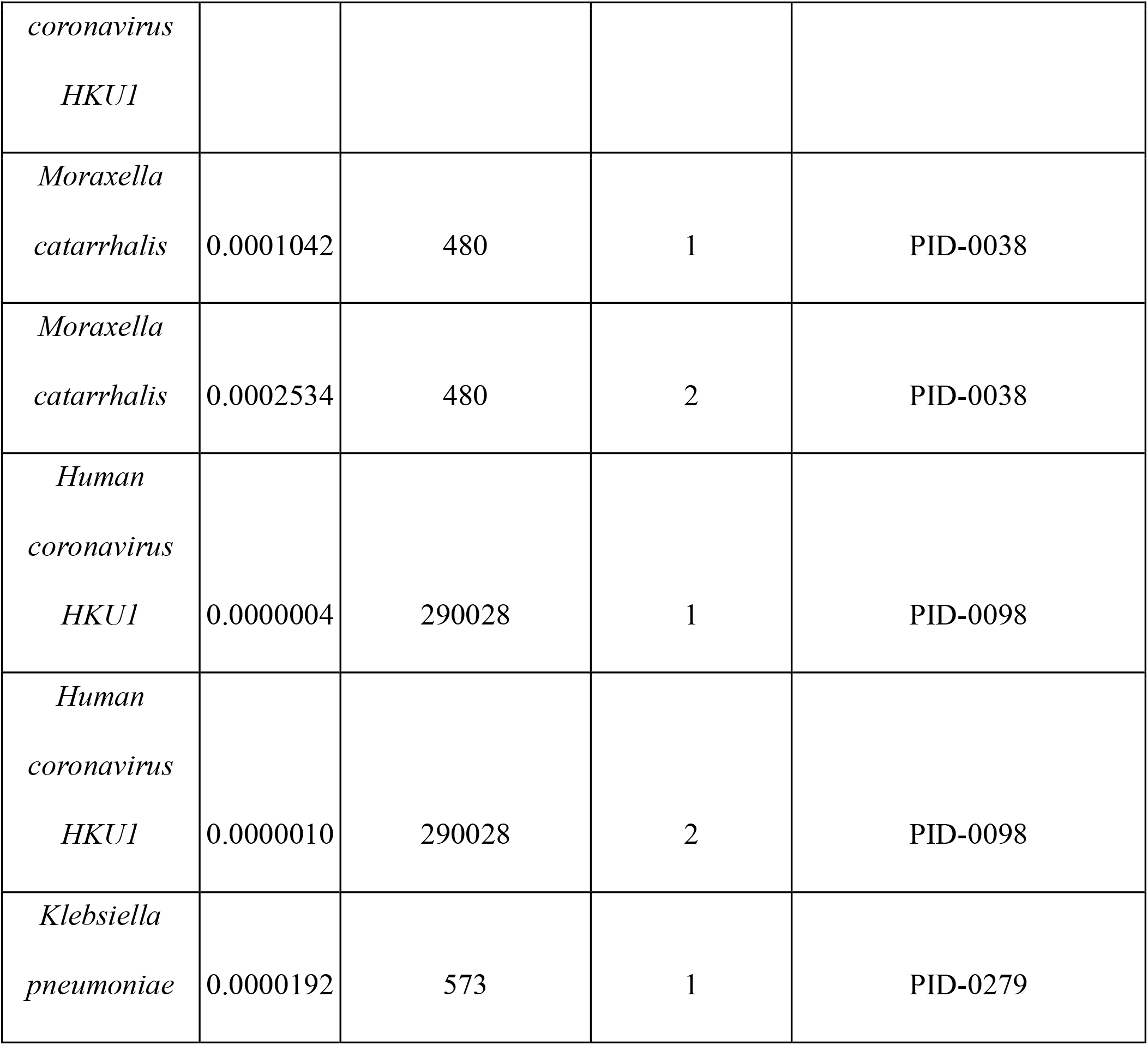
Significantly different abundance of pathogenic microorganisms between any sick time point and all healthy time points based on two sample t-test.

### Laboratory analysis

Throat samples were lysed by bead beating in RPB, as previously described (Hatch et al., 2019; Toma et al., 2020). Briefly, the samples were lysed using a combination of chemical and mechanical (bead beading) methods. RNA was extracted using silica beads and a series of washes, followed by elution in molecular biology grade water. DNA was degraded using RNase-free DNase. Prokaryotic and human rRNAs were removed using a subtractive hybridization method previously described (Hatch et al., 2019). Biotinylated DNA probes with sequences complementary to microbial and human rRNAs were added to total RNA, the mixture was heated and cooled, and the probe-rRNA complexes were removed using magnetic streptavidin beads. The remaining RNAs that contain an enriched mRNA fraction were converted to directional sequencing libraries using unique dual-barcoded adapters and ultrapure reagents. Libraries were pooled and quality controlled with dsDNA Qubit (Thermo Fisher Scientific) and Fragment Analyzer (Advanced Analytical) methods. Library pools were sequenced on Illumina NovaSeq instruments using 300 cycle kits.

### Bioinformatic analysis

Viome’s bioinformatics methods include quality control, strain-level taxonomic classification, and microbial gene expression. The quality control includes per-sample and per-batch quality metrics, such as the level of barcode hopping, batch contamination, positive and negative process controls, DNase efficacy, and the number of reads obtained per sample. Following the quality control, the paired-end reads are aligned to a catalog containing ribosomal RNA (rRNA), human genome, and 53,660 genomes spanning archaea, bacteria, fungi, protozoa, and viruses. Reads that map to rRNA or host are filtered out. Strain-level relative activities were computed from mapped reads via the expectation-maximization (EM) algorithm (Dempster et al., 1977). Relative activities at other levels of the taxonomic tree are then computed by aggregating according to taxonomic rank. Relative activities for the biological functions are computed by mapping paired-end reads to a catalog of 52,324,420 genes, quantifying gene-level relative activity with the EM algorithm, and then aggregating gene-level activity by KEGG Ortholog (KO) annotation (Kanehisa and Goto, 2000). The identified and quantified active microbial species and KOs for each sample are then used for downstream analysis.

For the identification of antimicrobial resistance genes, sequencing reads from the throat swab samples were mapped to a total of 5,735 NCBI antimicrobial resistance proteins using Diamond. Mappings with e-value > 1e-10 were discarded. For reads that were mapped to several targets, the mapping with the smallest e-value was kept.

### Data analysis

Statistical parameters, including transformations and significance, are reported in the figures and figure legends. To compare pairs of samples, we report Jaccard similarity (which ignores expression and considers overlap in genes detected), Spearman correlations (which are invariant to absolute expression levels of the genes and only consider the similarity of ranked expression), Pearson correlations on logged data (which measure the linear relationship between gene expression levels), and Hellinger distance (an appropriate distance measure for compositional data). Pearson and Spearman correlation coefficients were performed on centered log-ratio (CLR) transformed data (Aitchison, 1982), as is commonly done to reduce false discoveries due to the compositional nature of sequencing data. Statistical analyses were performed in python.

## Results

### Sequencing Metrics

Of the thirty-six throat samples that were analyzed from the six donors, the average number of reads per sample was 9,542,332 reads (standard deviation: 2,268,830 reads) and the average species richness was 479 species (standard deviation: 72 species). The top ten microbial species by abundance that were detected across all samples accounted for 29% of all microbial reads. The species were *Streptococcus salivarius* (accounting for 5.7% of microbial reads and detected in 100% of samples), *Veillonella atypica* (accounting for 4.9% of microbial reads and detected in 100% of samples), *Tannerella sp. oral taxon HOT-286* (accounting for 3.5% of microbial reads and detected in 88.9% of samples), *Gemella sanguinis* (accounting for 2.6% of microbial reads and detected in 100% of samples), *Veillonella sp. oral taxon 158* (accounting for 2.6% of microbial reads and detected in 100% of samples), *Prevotella shahii* (accounting for 2.4% of microbial reads and detected in 88.9% of samples), *Leptotrichia wadei* (accounting for 2.2% of microbial reads and detected in 100% of samples), *Streptococcus mitis* (accounting for 1.7% of microbial reads and detected in 100% of samples), *Streptococcus parasanguinis* (accounting for 1.7% of microbial reads and detected in 100% of samples), and *Streptococcus infantis* (accounting for 1.6% of microbial reads and detected in 100% of samples). Based on a post-hoc test with Mann-Whitney, there was no significant difference between any of the participants in terms of reads or species richness (figure 1A and 1B). The average percent of sequencing reads that mapped to microbial species (microbial load) was 7.57% (standard deviation: 4.74%). Participant PID-0001 showed significantly increased microbial load compared to all other participants (p<0.05) (figure 1C).

**Figure 1:**
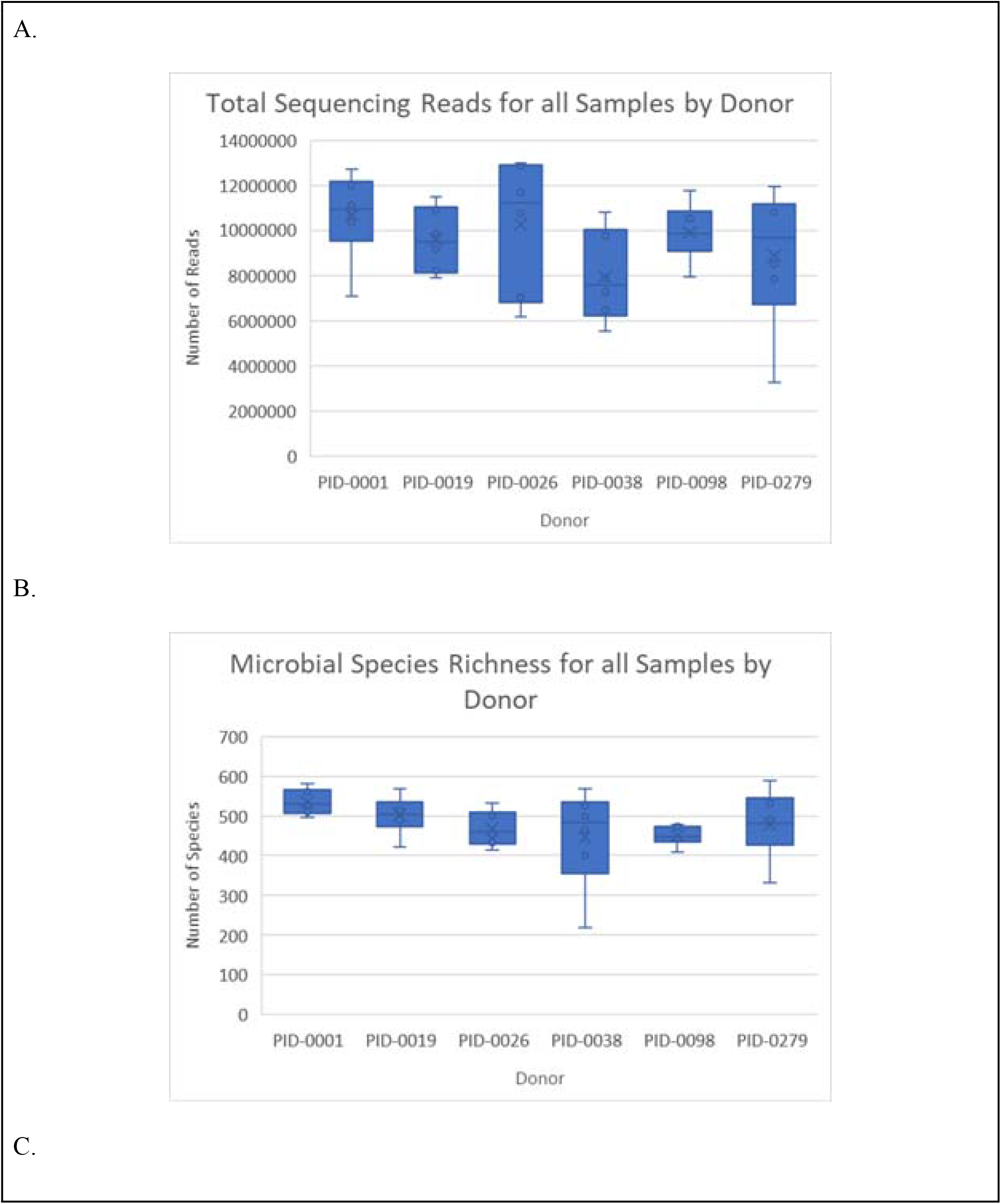
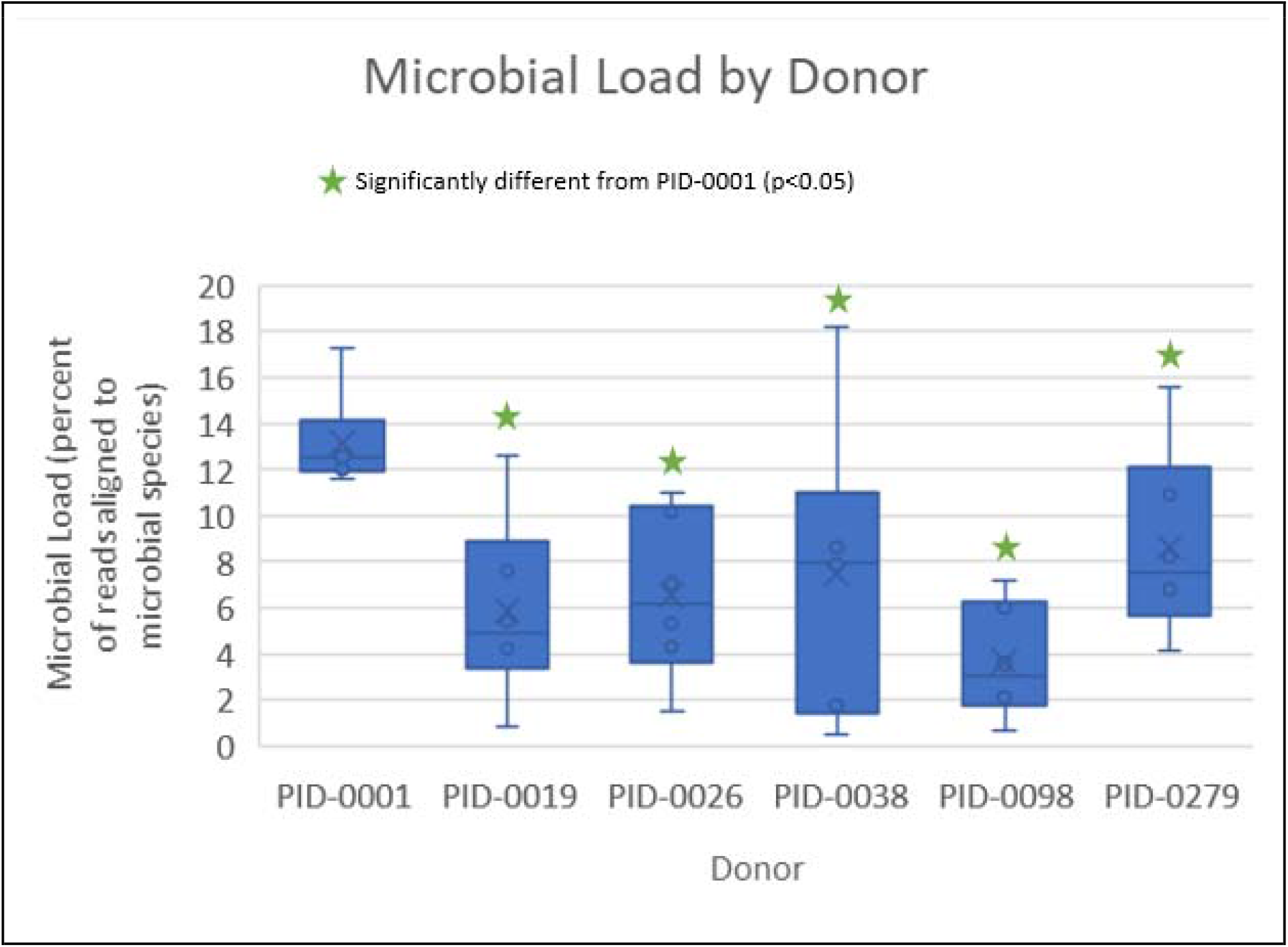
Comparison of sequencing metrics from all samples for all donors. Based on a post-hoc test with Mann-Whitney U (MWU) test, there is no significant difference between donors for (A) the number of sequencing reads and (B) species richness. PID-0001 has a significant elevation in (C) the microbial load of their samples. Each donor had six samples analyzed.

### Pathogen Detection and Characterization

Five of the six participants had a significant change in the relative abundance of one or more pathogenic organisms during the sickness compared to all of the intra-participant healthy samples (both pre and post-sickness) based on two sample t-test. Participants PID-0098 and PID-0026 had significant increases in *Human coronavirus HKU1* abundance, PID-0038 had a significant increase in *Moraxella catarrhalis* abundance, PID-00279 had a significant increase in *Klebsiella pneumoniae* abundance, and PID-0019 had significant increases in *Rhinovirus A, Streptococcus pneumoniae, and Haemophilus influenzae* abundance and a significant decrease in *Haemophilus parainfluenzae* abundance in the sick time points compared to all of the healthy time points (table 1, figure 2).

**Figure 2:**
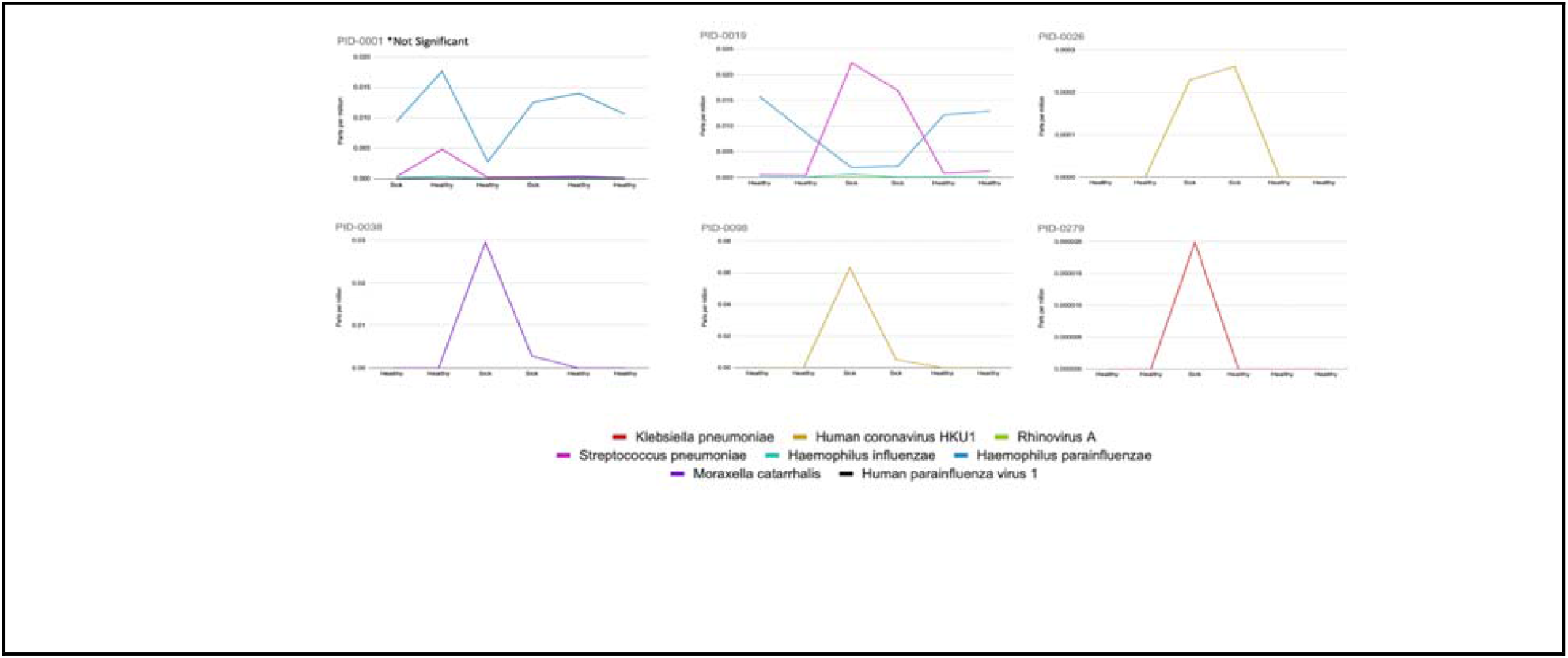
Significantly different relative abundances (in parts per million) of pathogenic microorganisms in throat samples during a sickness compared to all healthy timepoints based on two sample t-test. All participants showed at least one significantly different microorganism in response to the sickness, except for PID-0001.

Participant PID-0001 did not have a significant change in pathogenic microorganisms during the sickness (figure 2). PID-0001 primarily reported a severe cough which they have noted is a chronic cough that has afflicted them for over ten years. Their symptoms are consistent with that of persistent bacterial bronchitis in adults (Finch et al., 2019). The significantly higher microbial load (figure 1) observed in all of the throat swabs from PID-0001 compared to all other participants corroborates this probable diagnosis (Sibila et al., 2019). In addition, all samples from PID-0001 show the presence of *Haemophilus influenzae*, whose presence is associated with persistent bacterial bronchitis (Gallucci et al., 2020). These results indicate that the participant may be suffering from bronchiectasis characterized by a general overgrowth of microorganisms in the throat leading to airway and epithelial damage which is the root cause of their cough, as opposed to being caused by a transient pathogenic infection (Martin and Harrison, 2015).

For several participants, the symptomatic data (supplemental table 2) that was collected also aligned with expected symptoms related to the purported infections. PID-0098 and PID-0026, for example, both contracted *Human coronavirus HKU1* based on throat swab analysis. Their predominant symptom was severe rhinorrhea (runny nose) and mild sore throat. Rhinorrhea and sore throat are common symptoms of *Human coronavirus HKU1* infection (Kanwar et al., 2017). Based on throat microbiome analyses, PID-0019 had an infection dominated by *Streptococcus pneumoniae*. Their predominant complaints were a sore throat, body aches, and sinus pressure, which are consistent with symptoms observed in bacterial sinusitis, which *Streptococcus pneumoniae* is known to cause (CDC, 2019; Scheid and Hamm, 2004).

### Functional and Taxonomic Shifts in Response to the Sickness

In addition to the significant changes in infectious organisms in the sick conditions, three participants also showed significant changes in highly expressed microbial functions or highly abundant non-pathogenic microbial species between the sick and the healthy time points based on paired t-test and corrected using Benjamini-Hochberg FDR correction (figure 3). Several functions associated with carbohydrate metabolism (K00134 and K01624) were significantly upregulated in participants PID-0019 and PID-0279 during the sickness compared to after the sickness (p<0.05). In addition, the expression of elongation factor Tu (EF-Tu/K02358) was significantly elevated during the sickness compared to after the sickness for participant PID-0279 (p<0.05). Two participants also showed significant changes in the relative abundance of two commensal bacteria as a result of the sickness. PID-0038 showed a significant reduction in the abundance of *Prevotella sp. oral taxon 306* in response to the sickness (sick time point 1 p=0.0014, sick time point 2 p=0.0060). PID-0279 showed a significant reduction in the abundance of *Actinomyces sp. Marseille-P2825* in response to the sickness (p=0.0215). A metatranscriptomic analysis of the throat microbiome is able to provide additional information about the functional and community shifts observed in response to a sickness, which can provide further insights into disease progression.

**Figure 3:**
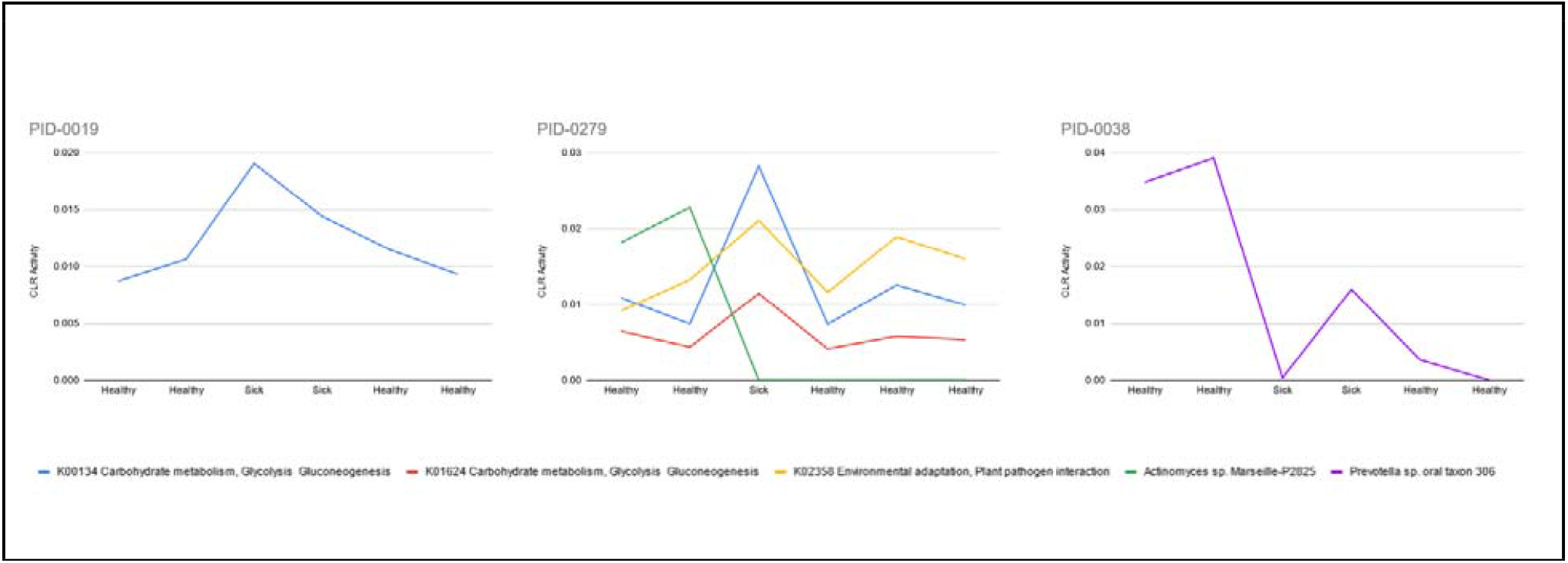
Significant changes in the expression of microbial functions and the abundance of commensal microorganisms in response to a sickness based on paired t-test and corrected using Benjamini-Hochberg FDR correction.

### Antibiotic Resistance Genes

A robust understanding of antibiotic resistance genes is critical for the proper treatment of bacterial infections. The improper use of antibiotics can result in harmful effects to the host (via off-target effects on human microbiomes) and can also exacerbate antibiotic resistance (Hayhoe et al., 2018). Metatranscriptomic analyses allow for the identification of the expressed antibiotic resistance genes to inform treatment options and the selection of effective antibiotics. In the dataset, 170 antibiotic resistance genes were identified, which could be used in a clinical setting to inform the treatment of the infection (figure 4, supplemental table 3).

**Figure 4:**
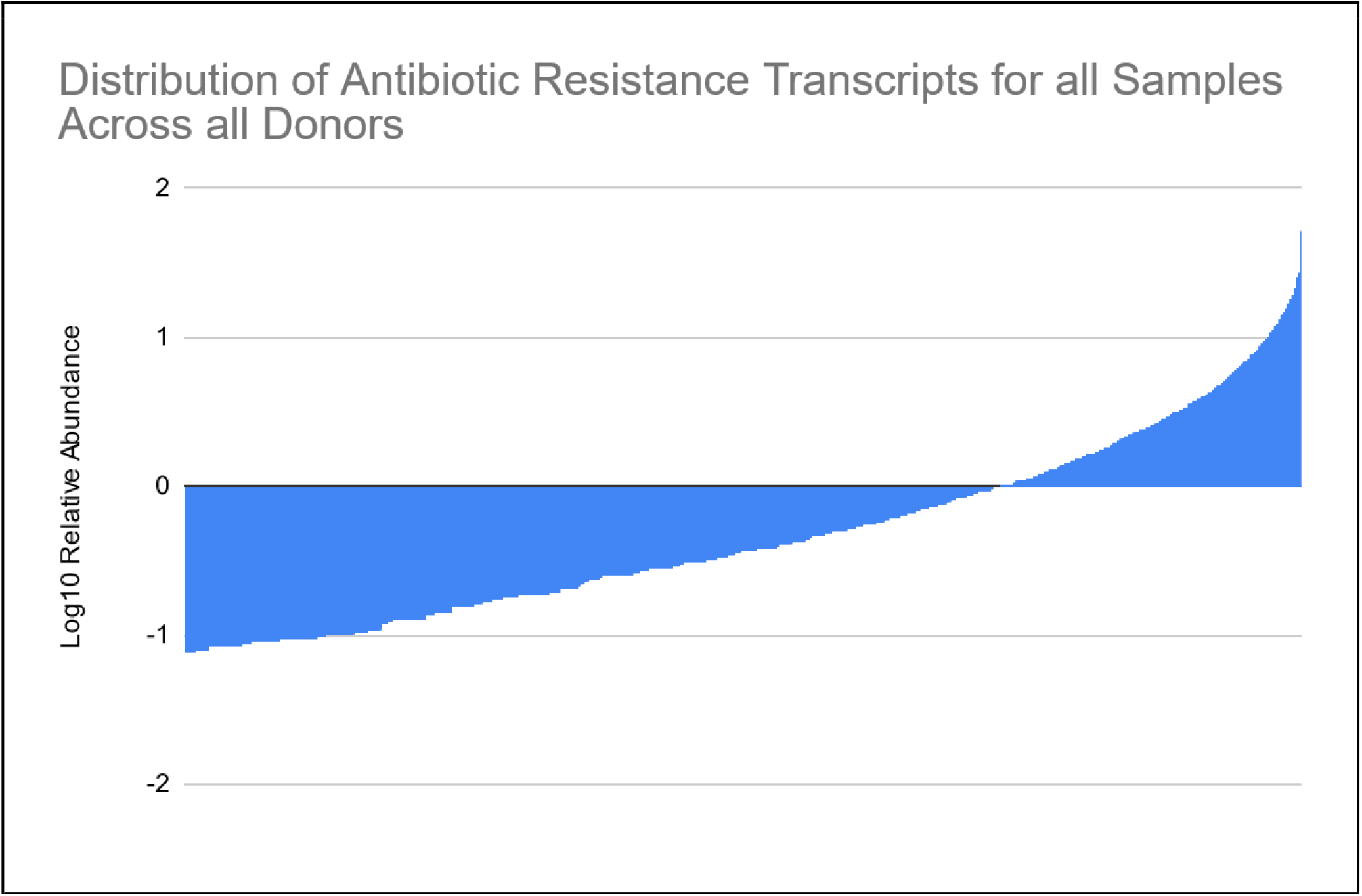
Expression level (log10 parts per million) distribution of all antibiotic resistance genes detected in all samples.

## Discussion

The current standard of care for the diagnosis of infectious diseases is inadequate and antiquated. Metatranscriptomic analysis of clinical samples presents an important advancement that allows for unbiased detection of all pathogens coupled with informative gene expression analysis (such as antibiotic resistance genes) that can inform treatment options. The throat microbiome can be readily sampled and can provide informative data on pathological biomarkers.

The metatranscriptomic method reported here is novel in that it is able to unbiasedly identify all pathogenic microorganisms at the highest resolution (strain level), can be used to investigate new and emerging pathogens, and can be deployed globally all while providing clinically relevant and actionable information in a timely manner. After physically removing the microbial and human rRNAs from the total RNA pool, the full length of all other RNAs are sequenced. Using the sequence information, accurate strain-level taxonomic classification can be performed for all microorganisms, including bacteria, fungi/molds, and RNA viruses, such as SARS-CoV-2, Influenza, etc. This approach will transform infectious disease diagnostics since healthcare providers will not need to rely on the symptoms to guess which test to run.

One of the most important advancements that metatranscriptomic analysis offers over traditional methodologies is its ability to identify pathogens that are difficult to detect with standard methods. For example, in the data presented in this paper, participant PID-0038 showed a likely infection with *Moraxella catarrhalis* (Table 1, figure 2). *Moraxella catarrhalis* is a common human pathogen that has been associated with several different diseases including sinusitis, ear infection, and COPD (Goldstein et al., 2009). Despite the prevalence of associated diseases with *Moraxella catarrhalis*, it was not recognized as a respiratory tract pathogen until recently, due to inadequate diagnostic methods and the inability of current diagnostic methods to differentiate *Moraxella catarrhalis* from other microorganisms such as *Neisseria* species (Goldstein et al., 2009). The methods reported in this paper overcome these limitations, allowing for unbiased identification and characterization of pathogens.

In this paper, it was also identified that participant PID-0001 likely suffers from chronic bronchiectasis, as opposed to a specific transient infection. Metatranscriptomic analysis of the entire throat microbiome is better able to differentiate between chronic conditions and infectious diseases with similar symptoms, which can facilitate proper treatment options.

Metatranscriptomics’ ability to quantify gene expression also offers exciting potential for the further characterization of infectious diseases and their role on host physiology, and for the development of more personalized treatment options. In the data presented in this paper, two participants showed alterations to the gene expression of two specific microbial functions in response to the sickness (figure 3). Increases in both carbohydrate metabolism and the expression of EF-Tu were observed. Participant PID-0279 showed increased EF-Tu expression which has been associated with the adhesion of pathogens to host cells and may assist in the pathogenicity of the infection (Harvey et al., 2019). Our findings also support previous research that has observed a co-occurrence between *Klebsiella pneumoniae* infection and a release of outer membrane vesicles containing EF-Tu (Harvey et al., 2019; Lee et al., 2012). Participants PID-0279 and PID-0019 also showed significantly increased functions associated with carbohydrate metabolism during the sickness. Modulations to carbohydrate metabolism pathways are important factors that allow pathogens to adapt to, and thrive in, different host niches, improving the overall success of the infection (Echlin et al., 2020). Finally, functional analyses are able to identify antibiotic resistance genes, allowing for informed treatment options that are best suited for the specific infection. Metatranscriptomic analysis is, therefore, able to provide a plethora of functional information which can add to an understanding of the disease progression, pathogenicity, and potential treatments.

Since the throat microbiome is an effective proxy to the lung microbiome, analyses of the throat microbiome also offer the opportunity to investigate several chronic conditions that are associated both with the throat and lung microbiomes. Throat sampling is easy to perform and could help to dramatically increase the understanding of how the throat and lung microbiomes impact human health and disease.

Although there are a myriad of benefits to the use of metatranscriptomics in infectious disease diagnostics, the current method does have several limitations. The primary limitation to the method is the complexity of the laboratory analyses. Although the process is automated, the laboratory space, reagent storage requirements, and training are limitations to implementing in diverse testing environments, for example in point of care settings. A limitation of this study is the limited sample size; to gain a robust understanding of the utility of metatranscriptomics in infectious disease diagnostics, further experiments should be performed.

Here we describe a novel throat metatranscriptome test that is capable of producing high-quality microbial taxonomic classifications and microbial gene expression profiles to facilitate the identification and characterization of pathogens. The test is also able to provide insight into the throat microbiome to further research its role in human health and disease. The sample collection described in this paper can be performed by anyone, anywhere, and samples can be stored and shipped at ambient temperature. The method is automatable, inexpensive, high throughput, and can be implemented in clinics and/or large-scale population studies.

## Data Availability

All data produced in the present study are available upon reasonable request to the authors

## Author Contributions

M Vuyisich and R Toma conceived and designed the methods. R Toma and N Duval performed sample and data collection. P Torres and F Camacho developed the bioinformatics pipeline. N Shen, J Chen, O Ogundijo, and G Banavar performed data analysis. All authors contributed to data interpretation. R Toma and N Duval contributed to the writing of the manuscript.

## Ethical conduct of research

The authors state that they have obtained appropriate institutional review board approval or have followed the principles outlined in the Declaration of Helsinki for all human or animal experimental investigations. In addition, for investigations involving human subjects, informed consent has been obtained from the participants involved.

## Financial and competing interests disclosure

All authors are current or former employees of Viome, Inc. The authors have no other relevant affiliations or financial involvement with any organization or entity with a financial interest in or financial conflict with the subject matter or materials discussed in the manuscript apart from those disclosed.

No writing assistance was utilized in the production of this manuscript.

## Notes

### Funding Statement

This study was funded by Viome Life Sciences

### Author Declarations

Viome IRB of Viome Life Sciences gave ethical approval for this work

## References

Aitchison J. The Statistical Analysis of Compositional Data. J Royal Statistical Soc Ser B Methodol 1982;44:139–60. https://doi.org/10.1111/j.2517-6161.1982.tb01195.x.

Bashiardes S, Zilberman-Schapira G, Elinav E. Use of Metatranscriptomics in Microbiome Research. Bioinform Biology Insights 2016;10:BBI.S34610. https://doi.org/10.4137/bbi.s34610.

Boutin S, Graeber SY, Weitnauer M, Panitz J, Stahl M, Clausznitzer D, et al. Comparison of Microbiomes from Different Niches of Upper and Lower Airways in Children and Adolescents with Cystic Fibrosis. Plos One 2015;10:e0116029. https://doi.org/10.1371/journal.pone.0116029.

Caliendo AM, Gilbert DN, Ginocchio CC, Hanson KE, May L, Quinn TC, et al. Better Tests, Better Care: Improved Diagnostics for Infectious Diseases. Clin Infect Dis Official Publ Infect Dis Soc Am 2013;57:S139–70. https://doi.org/10.1093/cid/cit578.

Castro-Nallar E, Bendall ML, Pérez-Losada M, Sabuncyan S, Severance EG, Dickerson FB, et al. Composition, taxonomy and functional diversity of the oropharynx microbiome in individuals with schizophrenia and controls. Peerj 2015;3:e1140. https://doi.org/10.7717/peerj.1140.

CDC. Sinus Infection (Sinusitis) | Antibiotic Use | CDC. Antibiotic Prescribing and Use 2019. https://www.cdc.gov/antibiotic-use/sinus-infection.html?CDC_AA_refVal=https%3A%2F%2Fwww.cdc.gov%2Fantibiotic-use%2Fcommunity%2Ffor-patients%2Fcommon-illnesses%2Fsinus-infection.html (accessed July 28, 2021).

Dempster AP, Laird NM, Rubin DB. Maximum Likelihood from Incomplete Data Via the EM Algorithm. Journal of the Royal Statistical Society: Series B (Methodological) 1977;39:1–22.

Echlin H, Frank M, Rock C, Rosch JW. Role of the pyruvate metabolic network on carbohydrate metabolism and virulence in Streptococcus pneumoniae. Mol Microbiol 2020;114:536–52. https://doi.org/10.1111/mmi.14557.

Finch S, Carreto L, Hani A-L, Browne A, Fardon T, Chalmers J. Persistent bacterial bronchitis in adults – a precursor to Bronchiectasis? Adv Exp Med Biol 2019:PA4585. https://doi.org/10.1183/13993003.congress-2019.pa4585.

Gallucci M, Pedretti M, Giannetti A, Palmo E di, Bertelli L, Pession A, et al. When the Cough Does Not Improve: A Review on Protracted Bacterial Bronchitis in Children. Frontiers Pediatrics 2020;8:433. https://doi.org/10.3389/fped.2020.00433.

Goldstein EJC, Murphy TF, Parameswaran GI. Moraxella catarrhalis, a Human Respiratory Tract Pathogen. Clin Infect Dis 2009;49:124–31. https://doi.org/10.1086/599375.

Gong H, Shi Y, Zhou X, Wu C, Cao P, Xu C, et al. Microbiota in the Throat and Risk Factors for Laryngeal Carcinoma. Appl Environ Microb 2014;80:7356–63. https://doi.org/10.1128/aem.02329-14.

Gong H-L, Shi Y, Zhou L, Wu C-P, Cao P-Y, Tao L, et al. The Composition of Microbiome in Larynx and the Throat Biodiversity between Laryngeal Squamous Cell Carcinoma Patients and Control Population. Plos One 2013;8:e66476. https://doi.org/10.1371/journal.pone.0066476.

Harvey KL, Jarocki VM, Charles IG, Djordjevic SP. The Diverse Functional Roles of Elongation Factor Tu (EF-Tu) in Microbial Pathogenesis. Front Microbiol 2019;10:2351. https://doi.org/10.3389/fmicb.2019.02351.

Hatch A, Horne J, Toma R, Twibell BL, Somerville KM, Pelle B, et al. A Robust Metatranscriptomic Technology for Population-Scale Studies of Diet, Gut Microbiome, and Human Health. Int J Genomics 2019;2019:1718741. https://doi.org/10.1155/2019/1718741.

Hayhoe B, Butler CC, Majeed A, Saxena S. Telling the truth about antibiotics: benefits, harms and moral duty in prescribing for children in primary care. J Antimicrob Chemoth 2018;73:2298–304. https://doi.org/10.1093/jac/dky223.

He S, Wurtzel O, Singh K, Froula JL, Yilmaz S, Tringe SG, et al. Validation of two ribosomal RNA removal methods for microbial metatranscriptomics. Nat Methods 2010;7:807–12. https://doi.org/10.1038/nmeth.1507.

Ibironke O, McGuinness LR, Lu S-E, Wang Y, Hussain S, Weisel CP, et al. Species-level evaluation of the human respiratory microbiome. Gigascience 2020;9. https://doi.org/10.1093/gigascience/giaa038.

Kanehisa M, Goto S. KEGG: kyoto encyclopedia of genes and genomes. Nucleic Acids Res 2000;28:27–30. https://doi.org/10.1093/nar/28.1.27.

Kanwar A, Selvaraju S, Esper F. Human Coronavirus (CoV)-HKU1 Infection among Adults in Cleveland, OhioCoV-HKU1 in Adults in Cleveland, Ohio. Open Forum Infect Dis 2017;4:ofx052.. https://doi.org/10.1093/ofid/ofx052.

Knight R, Jansson J, Field D, Fierer N, Desai N, Fuhrman JA, et al. Unlocking the potential of metagenomics through replicated experimental design. Nat Biotechnol 2012;30:513–20. https://doi.org/10.1038/nbt.2235.

Larremore DB, Wilder B, Lester E, Shehata S, Burke JM, Hay JA, et al. Test sensitivity is secondary to frequency and turnaround time for COVID-19 surveillance. Medrxiv 2020:2020.06.22.20136309. https://doi.org/10.1101/2020.06.22.20136309.

Lee JC, Lee EJ, Lee JH, Jun SH, Choi CW, Kim SI, et al. Klebsiella pneumoniae secretes outer membrane vesicles that induce the innate immune response. Fems Microbiol Lett 2012;331:17– 24. https://doi.org/10.1111/j.1574-6968.2012.02549.x.

Lee KH, Gordon A, Shedden K, Kuan G, Ng S, Balmaseda A, et al. The respiratory microbiome and susceptibility to influenza virus infection. Plos One 2019;14:e0207898. https://doi.org/10.1371/journal.pone.0207898.

Mao Q, Jiang F, Yin R, Wang J, Xia W, Dong G, et al. Interplay between the lung microbiome and lung cancer. Cancer Lett 2018;415:40–8. https://doi.org/10.1016/j.canlet.2017.11.036.

Martin MJ, Harrison TW. Causes of chronic productive cough: An approach to management. Resp Med 2015;109:1105–13. https://doi.org/10.1016/j.rmed.2015.05.020.

Miller S, Chiu C, Rodino KG, Miller MB. Point-Counterpoint: Should We Be Performing Metagenomic Next-Generation Sequencing for Infectious Disease Diagnosis in the Clinical Laboratory? J Clin Microbiol 2019;58. https://doi.org/10.1128/jcm.01739-19.

Moffatt MF, Cookson WO. The lung microbiome in health and disease. Clin Med 2017;17:525– 9. https://doi.org/10.7861/clinmedicine.17-6-525.

Scheid DC, Hamm RM. Acute bacterial rhinosinusitis in adults: part I. Evaluation. Am Fam Physician 2004;70:1685–92.

Sibila O, Laserna E, Shoemark A, Keir HR, Finch S, Rodrigo-Troyano A, et al. Airway Bacterial Load and Inhaled Antibiotic Response in Bronchiectasis. Am J Resp Crit Care 2019;200:33–41. https://doi.org/10.1164/rccm.201809-1651oc.

Toma R, Pelle B, Duval N, Parks MM, Gopu V, Tily H, et al. A clinically validated human capillary blood transcriptome test for global systems biology studies. Biorxiv 2020:2020.05.22.110080. https://doi.org/10.1101/2020.05.22.110080.

Tsang TK, Lee KH, Foxman B, Balmaseda A, Gresh L, Sanchez N, et al. Association Between the Respiratory Microbiome and Susceptibility to Influenza Virus Infection. Clin Infect Dis 2019;71:1195–203. https://doi.org/10.1093/cid/ciz968.

Wylezich C, Papa A, Beer M, Höper D. A Versatile Sample Processing Workflow for Metagenomic Pathogen Detection. Sci Rep-Uk 2018;8:13108. https://doi.org/10.1038/s41598-018-31496-1.

